# B cell immune repertoire sequencing in tobacco cigarette smoking, vaping, and chronic obstructive pulmonary disease in the COPDGene cohort

**DOI:** 10.1101/2024.10.07.24315038

**Authors:** Matthew Moll, Zhonghui Xu, Adel Boueiz, Min Hyung Ryu, Edwin K. Silverman, Michael H. Cho, Craig P. Hersh, Maor Sauler, Francesca Polverino, Gregory L. Kinney, Jeffrey L. Curtis, Laura E. Crotty-Alexander, Christopher Vollmers, Peter J. Castaldi

## Abstract

**Rationale:** Cigarette smoking (CS) impairs B cell function and antibody production, increasing infection risk. The impact of e-cigarette use (’vaping’) and combined CS and vaping (’dual-use’) on B cell activity is unclear.

**Objective:** To examine B cell receptor sequencing (BCR-seq) profiles associated with CS, vaping, dual-use, COPD-related outcomes, and demographic factors.

**Methods:** BCR-seq was performed on blood RNA samples from 234 participants in the COPDGene study. We assessed multivariable associations of B cell function measures (immunoglobulin heavy chain (IGH) subclass expression and usage, class-switching, V-segment usage, and clonal expansion) with CS, vaping, dual-use, COPD severity, age, sex, and race. We adjusted for multiple comparisons using the Benjamini-Hochberg method, identifying significant associations at 5% FDR and suggestive associations at 10% FDR.

**Results:** Among 234 non-Hispanic white (NHW) and African American (AA) participants, CS and dual-use were significantly positively associated with increased secretory IgA production, with dual-use showing the strongest associations. Dual-use was positively associated with class switching and B cell clonal expansion, indicating increased B cell activation, with similar trends in those only smoking or only vaping. We observed significant associations between race and IgG antibody usage. AA participants had higher IgG subclass proportions and lower IgM usage compared to NHW participants.

**Conclusions:** CS and vaping additively enhance B cell activation, most notably in dual-users. Self-reported race was strongly associated with IgG isotype usage. These findings highlight associations between B cell activation and antibody transcription, suggesting potential differences in immune and vaccine responses linked to CS, vaping, and race.

## Introduction

Use of electronic cigarettes, i.e. vaping, has increased substantially since their introduction to the U.S. market in 2007^1^. Numerous studies have demonstrated that vaping induces inflammatory responses and has adverse health effects^2–6^. More precise characterization of the inflammatory effects of vaping may better define its effects on health, both for vaping alone and vaping in conjunction with combustible cigarettes, i.e. dual-use.

B cells participate in adaptive immunity largely by producing antibodies that protect mucosal surfaces and provide antigen-specific responses to infection. Combustible tobacco cigarette smoking (CS) has adverse effects on B cells resulting in increased susceptibility to infections^7^. Proper B cell function depends on B cell activation, a process in which naïve B cells are activated by exposure to an antigen. This process triggers clonal expansion of B-cell populations with specifically rearranged B-cell receptor (BCR) genes, which encode the specific immunoglobulin (Ig) produced by each clone. In conjunction with T-cell help, these activated clones undergo somatic hypermutation, becoming optimized to bind specific antigens. BCR sequencing (BCR-seq) allows for the identification and sequence-specific characterization of B cells and expanded B cell clones, providing rich characterization of the B cell response^8^.

We hypothesized that vaping and dual-use alter B cell function and the ability of B cells to respond appropriately to antigens through activation and antibody production. To address this question, and to determine whether any of these B cell changes are associated with measures of t chronic obstructive pulmonary disease (COPD), we performed BCR-seq in 234 participants from the Genetic Epidemiology of COPD (COPDGene)^9^ Study, a large study of individuals who currently or previously smoked that is enriched for participants with COPD.

## Methods

### Study population

Written informed consent was obtained from all study participants, and institutional review board approval was obtained at all study centers. The Genetic Epidemiology of COPD (COPDGene)^9^ study enrolled 10,198 non-Hispanic white (NHW) and African American (AA) individuals who smoked 10 or more pack-years of cigarettes during their lifetime and who were aged 45-80 at study enrollment. COPDGene is an ongoing longitudinal study with completed enrollment, 5-year, and 10-year visits. At each visit, anthropometric measurements, spirometry, chest computed tomography (CT) imaging, and blood samples were collected. At the 5-year follow up visit, we collected questionnaire data on use of cigarette and e-cigarette products. All data in this paper comes from the 5-year study visit where both blood RNA-seq and vaping data are available.

### Participant selection

Selection of participants for BCR-seq^8^ was performed using a stratified random sampling approach as follows. First, all participants with available blood RNA and complete vaping and CS data from COPDGene Phase 2 at the time of participant selection were considered (n=3,601). They were stratified into five groups based on vaping and CS status – never smokers, former combustible cigarette smokers, current cigarette smokers (without current vaping), current vapers (without current cigarette use), and current dual-users (vaping and cigarette use). All participants in the current vaper (n=41) and dual-user group (n=57) with available samples were selected for BCR-seq, and the remaining 136 participants were randomly sampled from the set of available participants in the other CS groups. Participants taking oral corticosteroids were excluded from the analysis as these medications are known to modify B cell function.

### Definition of key study variables

CS and vaping behavior were ascertained by self-report. Vapers were participants who reported using at least one e-cigarette within the prior week and had a history of smoking tobacco cigarettes, but not within the last 30 days. Current cigarette users reported current smoking with an average of at least one cigarette per day without any e-cigarette use. Dual-users were vapers who also reported current CS, and former cigarette users were defined as those who reported a history of smoking but did not meet criteria for current CS or vaping. In most of the reported analyses, former cigarette users are used as the reference group.

Information on the age, sex, and race of participants was elicited through self-report. For sex, participants were asked if they were male or female. For race, participants were asked if they were NHW, Black or African American, Asian, Pacific Islander, American Indian or Alaska Native, or Other with the option to select multiple categories. By design, inclusion in COPDGene was limited to participants self-identifying as NHW or AA. A separate ethnicity question asked participants if they were Hispanic or Latino. For this study, participants were coded as NHW if they indicated “White” and AA if they indicated “Black or African American.”

In the U.S., COPD primarily develops in the setting of cigarette smoking exposure, and B cell lymphoid follicles are associated with COPD severity^10–12^. Therefore, we examined the association of BCR-seq measures with COPD and COPD-related traits. COPD status was determined by GOLD spirometry grades based on post-bronchodilator spirometry testing where participants were grouped into normal spirometry (FEV1/FVC ≥ 0.7 and FEV1 % predicted ≥ 80%) or GOLD spirometry grade 1, GOLD grade 2-4, or preserved ratio with impaired spirometry (PRISm)^13^. For all analyses, the reference group included formerly smoking individuals with normal spirometry. Global Lung Initiative (GLI) race-neutral equations were used to calculate % predicted spirometry values. Computed tomography (CT) imaging measures of emphysema and airway wall thickness were generated by Thirona (https://thirona.eu/) and the following measures were analyzed: % low attenuation area less than −950 Hounsfield units for emphysema and airway wall thickness as % of overall airway volume (wall area percent^14^).

### B-cell receptor sequencing library preparation

Details regarding generation of RNAseq data in COPDGene were previously published^15^. Whole blood was collected and stored in PAXgene Blood RNA tubes, and total RNA was extracted using Qiagen PreAnalytiX PAXgene Blood miRNA Kit (Qiagen, Valencia, CA). Sequencing libraries were prepared using 200 ng of total RNA as input following a protocol modified from ^8^. Additional details regarding library preparation and data processing can be found in the Supplementary Methods.

We generated adaptive immune receptor repertoire sequencing data for B cell receptors (hereafter, ‘BCR-seq’) data using a set of isotype-specific immunoglobulin heavy chain (IGH) constant region primers. Reads were aligned to International Immunogenetics Information System (IMGT) reference germline sequences, and clonal relationships between BCR sequences were inferred using the spectralClones function from the scoper R package contained within the Immcantation suite of software packages (https://immcantation.readthedocs.io/en/stable/about.html). Mutated sequences were defined as sequences that were aligned but differed from the IMGT reference by one or more bases.

Uniquely identified BCR sequences were quantified to represent antibody isotype expression (log 2 counts of the number of unique BCR sequences present within each isotype class) and usage (number of unique BCR sequences present within each isotype class divided by the total number of BCR sequences), B cell activation measured through class switching (number of unique BCR sequences in the IgA, IgG, and IgE isotypes divided by the total number of BCR sequences), length of the CDR3 region in nucleotides, and the clonal diversity of the B cell population in each individual as measured by Hill numbers^16^. V-segment usage was defined as the number of unique and mutated BCR sequences containing a specific V-segment (as defined by IGHV genes from the IMGT reference) divided by total number of unique BCR sequences. For each BCR-seq measure, we analyzed only those measures where the isotype or V-segment class in question was present at >1% of the total unique sequences for 25% of the participants or more. The one exception was measurement of the IgE isotype which was analyzed despite being below this threshold due to its established clinical importance.

### B-cell Receptor Sequencing Measures

BCR sequencing involves sequencing transcripts of the B cell receptor using a set of primers targeting the Fc-region of the immunoglobin heavy chain (IGH). This provides comprehensive assessment of the BCR repertoire including antibody isotype (IgM, IgD, IgA, IgG, and IgE), V-segments corresponding to the variable region that determines antibody specificity, and clonal expansion and somatic hypermutation of specific B cell populations. These sequence counts are summarized into quantitative measures of 1) isotype usage (proportion of antibody transcripts for each isotype within each individual), 2) isotype expression (log2 transformed counts that represent the number of unique B cells per isotype within each individual), 3) class switching (proportion of class-switched B cells per individual), 4) V-segment usage (proportion of antibody transcripts for each V-segment within each individual), 5) CDR3 length by isotype, and 6) B cell clonal diversity measured by Hill numbers. For duplicated sequences, we only counted the sequence once, which means our measures represent numbers of B cells rather than number of transcripts.

### Statistical Analysis

We performed analyses in R version >4.0 (www.r-project.org). We assessed normality of continuous variables by visual inspection of histograms. Results are shown as mean ± standard deviation or median [interquartile range], as appropriate. Differences in continuous variables were assessed with Student t-tests or Wilcoxon tests. Categorical variables were compared by ANOVA or Kruskal-Wallis tests, as appropriate. We considered false discovery rate (FDR)-adjusted Benjamini-Hochberg^17^ p-values less below 0.05 to be significant and between 0.05 and 0.1 to be suggestive.

For each of the BCR-seq measures, we used univariable analysis and multivariable regression to examine the association between each of these measures with CS, vaping, and dual-use.

Further, we examined associations to age, sex, and race as well as COPD affection status (GOLD 2-4), CT emphysema measures ( % low attenuation area (LAA) < −950 Hounsfeld units (HU)^14^), and CT airway wall thickness (wall area %^14^).

For analyses of smoking/vaping, demographic variables, and GOLD spirometry grade, multivariable models included the following covariates: age, sex, self-identified race, vaping/smoking behavior, GOLD grade, pack-years of smoking, and inhaled corticosteroid use. For CT imaging measures, models were additionally adjusted for CT scanner model. To visualize the results, we constructed violin plots and heatmaps.

Sensitivity analyses were performed for the significant associations observed with smoking/vaping status and self-reported race adjusting for self-reported income level and social deprivation index, a measure of area-level deprivation^18^, and principal components of genetic ancestry. We additionally performed interaction analyses between self-reported race and smoking/vaping variables by including the main effects and cross-product interaction terms in a regression model.

## Results

### Characteristics of study participants

A schematic of our study design is shown in Figure 1. We included 234 COPDGene NHW and AA participants with smoking/vaping and BCR-seq data, and a table of their characteristics is shown in Table 1. Compared to other groups, dual-users were more likely to be younger, NHW, have more pack-years of cigarette smoking (CS), lower FEV_1_ % predicted, and thicker airway walls. Compared to dual-users, individuals who only vaped were slightly older, were less likely to be male, had similar pack-years of smoking, but had higher FEV_1_ % predicted, and more quantitative emphysema (% LAA < −950 HU).

**Figure 1:**
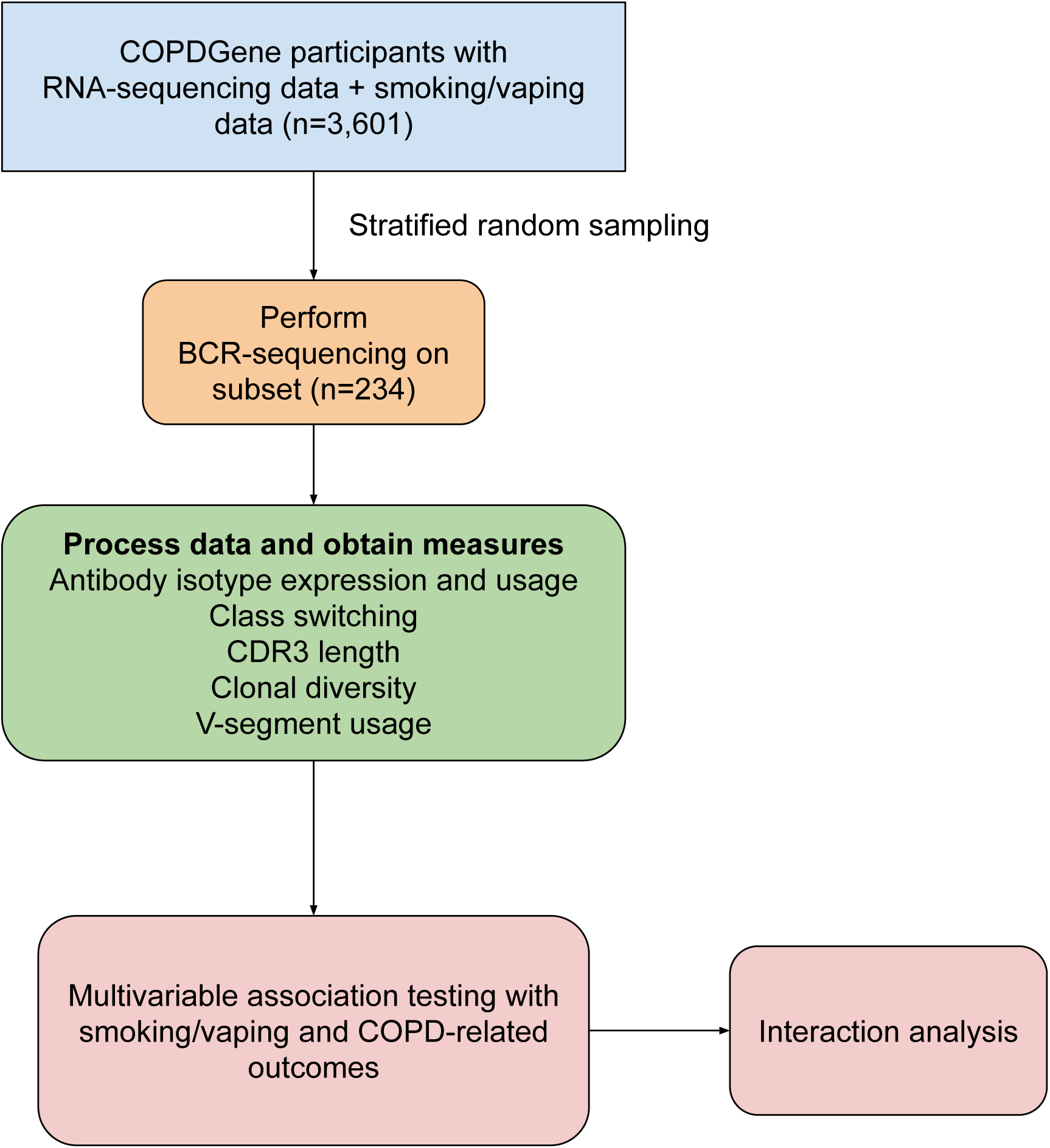
Schematic of study design. COPDGene = Genetic Epidemiology of COPD study. BCR = B cell receptor. COPD = chronic obstructive pulmonary disease.

**Table 1.** Characteristics of Study Participants.

| **Table 1. Characteristics of Study Participants** |  |  |  |  |  |  |  |
| --- | --- | --- | --- | --- | --- | --- | --- |
|  | **former smoker**<br>**(N=44)** | **never**<br>**(N=41)** | **vaping**<br>**(N=41)** | **cigarette**<br>**smoking**<br>**(N=51)** | **dual**<br>**(N=57)** | **overall**<br>**(N=234)** | **p-value** |
| age | 69.6 (7.42) | 65.3 (9.96) | 64.4 (6.56) | 62.0 (7.21) | 61.3 (6.47) | 64.2 (8.04) | <0.001 |
| sex |  |  |  |  |  |  |  |
| female | 23 (52.3%) | 29 (70.7%) | 26 (63.4%) | 31 (60.8%) | 29 (50.9%) | 138 (59.0%) | 0.403 |
| male | 21 (47.7%) | 12 (29.3%) | 15 (36.6%) | 20 (39.2%) | 28 (49.1%) | 96 (41.0%) |  |
| race |  |  |  |  |  |  |  |
| AA | 6 (13.6%) | 2 (4.9%) | 4 (9.8%) | 28 (54.9%) | 15 (26.3%) | 55 (23.5%) | <0.001 |
| NHW | 38 (86.4%) | 39 (95.1%) | 37 (90.2%) | 23 (45.1%) | 42 (73.7%) | 179 (76.5%) |  |
| cigarette pack-years | 43.0 (23.3) | 0 (0) | 52.2 (24.6) | 48.5 (27.8) | 52.9 (25.3) | 40.7 (29.8) | <0.001 |
| GOLD spirometry grade |  |  |  |  |  |  |  |
| Normal spirometry | 15 (34.1%) | 39 (95.1%) | 19 (46.3%) | 22 (43.1%) | 19 (33.3%) | 114 (48.7%) | <0.001 |
| GOLD 1 | 6 (13.6%) | 0 (0%) | 5 (12.2%) | 3 (5.9%) | 6 (10.5%) | 20 (8.5%) |  |
| GOLD 2,3,4 | 21 (47.7%) | 1 (2.4%) | 13 (31.7%) | 20 (39.2%) | 29 (50.9%) | 84 (35.9%) |  |
| PRISm | 2 (4.5%) | 1 (2.4%) | 4 (9.8%) | 6 (11.8%) | 3 (5.3%) | 16 (6.8%) |  |
| FEV1, % of predicted | 75.3 (27.7) | 74.1 (26.8) | 107 (13.2) | 72.8 (21.1) | 82.0 (24.9) | 81.3 (26.5) | <0.001 |
| FEV1/FVC | 0.633 (0.149) | 0.795 (0.0479) | 0.672 (0.142) | 0.688 (0.141) | 0.642 (0.158) | 0.682 (0.146) | <0.001 |
| CT emphysema | 6.30 (7.55) | 1.24 (1.44) | 3.43 (4.90) | 3.43 (7.13) | 3.10 (5.76) | 3.45 (5.99) | 0.002 |
| CT airway wall thickness | 51.1 (6.34) | 42.7 (4.81) | 48.4 (8.79) | 52.6 (8.97) | 52.6 (9.14) | 49.8 (8.67) | <0.001 |
| Values are mean (standard deviation) for continuous variables and N (%) for categorical variables. CT emphysema measurement is % low attenuation area <-950 Hounsfield units. CT airway wall thickness is defined as (area of segmental airway walls / overall airway area). FEV1 = forced expiratory volume in 1 second. FEV1/FVC = FEV1/forced vital capacity. PRISm = preserved ratio with impaired spirometry. AA= African American. NHW = non-Hispanic white. CT = computed tomography. Global Lung Initiative (GLI) race-neutral equations were used to calculate % predicted values. P-values test differences across all groups (analysis of variance (ANOVA)). |  |  |  |  |  |  |  |

### Associations between BCR-seq measures and cigarette smoking, vaping, and dual-use

Significant (q-value < 0.05) and suggestive (q-value <0.1) associations for vaping, CS, and dual-use are shown in Table 2 (Tables E1-E5 contain complete model results). Overall, we observed that the most pronounced changes in antibody production were associated with dual-use. Specifically, dual-use resulted in a shift in isotype usage towards IgA and away from IgM (Figure 2A). It was also associated with increased class switching suggestive of B cell activation (Figure 2B) and increased usage of specific V-segments. Clonality analysis also demonstrated reduced antibody diversity for participants engaged in dual-use (Figure E1), suggesting that there is a greater amount of B cell clonal expansion in this group. Since CS status is often associated with socioeconomic variables, we tested these associations after adjusting for income level and social deprivation index, a composite measure of area-level deprivation, which had minimal effect on the significance of these associations (Table E6).

**Figure 2.**
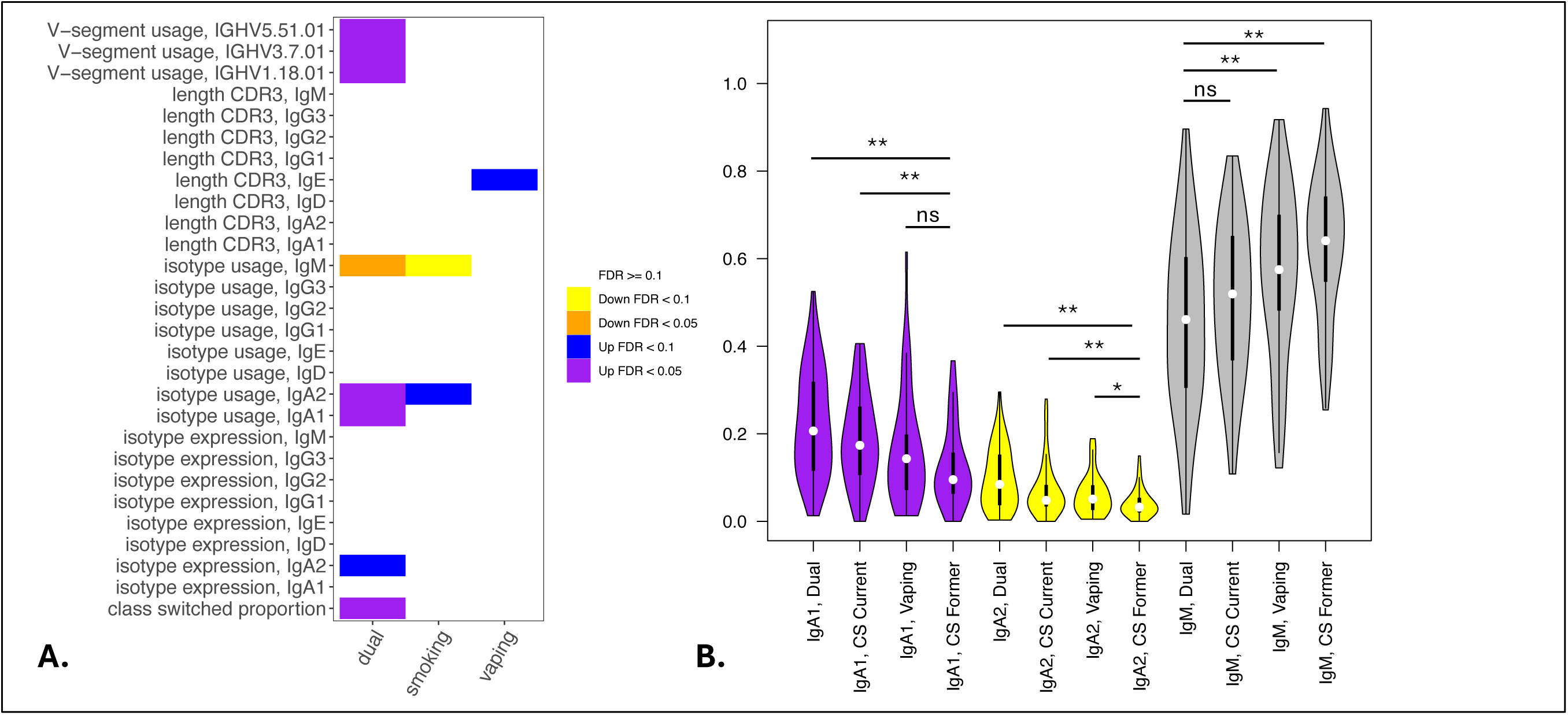
Associations of BCR Measures with Vaping and Cigarette Smoking. Significant associations between BCR measures and vaping, cigarette smoking, and dual-use from multivariable models analyzing class switching, isotype expression and usage, V-segment usage, and CDR3 length (in nucleotides) are shown in Panel A. Panel B shows IgA and IgM isotype usage among participants engaged in current cigarette smoking, vaping, or dual-use with former smokers included for comparison. Significance is assessed by t-tests. * p<=0.05, ** p<=0.005, ns p>0.05.

**Table 2.** Significant BCR associations to vaping, cigarette smoking, and dual-use.

| BCR measure | Dual-use |  |  | Cigarette smoking |  |  | Vaping |  |  |
| --- | --- | --- | --- | --- | --- | --- | --- | --- | --- |
| | $\beta$ (se) | P-value | q-value | $\beta$ (se) | P-value | q-value | $\beta$ (se) | P-value | q-value |
| IgA2 count | 0.879 (0.294) | 3.1E-03 | 0.05 | 0.766 (0.306) | 0.01 | 0.11 | 0.432 (0.301) | 0.15 | 0.48 |
| IgA1 usage | 0.095 (0.023) | 7.5E-05 | 2.7E-03 | 0.061 (0.025) | 0.01 | 0.11 | 0.051 (0.024) | 0.04 | 0.19 |
| IgA2 usage | 0.059 (0.011) | 2.8E-07 | 7.0E-05 | 0.035 (0.012) | 0.00 | 0.05 | 0.027 (0.012) | 0.02 | 0.12 |
| IgM usage | -0.173 (0.038) | 1.1E-05 | 8.9E-04 | -0.113 (0.04) | 0.01 | 0.07 | -0.087 (0.04) | 0.03 | 0.17 |
| class switching proportion | 0.111 (0.027) | 5.4E-05 | 2.6E-03 | 0.064 (0.028) | 0.02 | 0.14 | 0.05 (0.028) | 0.07 | 0.31 |
| IGHV1.18.01 usage | 0.005 (0.001) | 4.9E-04 | 0.01 | 0.004 (0.001) | 0.01 | 0.11 | 0.004 (0.001) | 0.02 | 0.12 |
| IGHV3.7.01 usage | 0.006 (0.002) | 3.2E-04 | 9.6E-03 | 0.003 (0.002) | 0.05 | 0.24 | 0.002 (0.002) | 0.28 | 0.62 |
| IGHV5.51.01 usage | 0.009 (0.002) | 6.2E-05 | 2.6E-03 | 0.005 (0.002) | 0.02 | 0.12 | 0.006 (0.002) | 0.01 | 0.11 |
| IgE CDR3 length | -0.384 (2.294) | 0.87 | 0.96 | 0.41 (2.392) | 0.86 | 0.96 | 6.755 (2.427) | 0.01 | 0.08 |
| Count values are log2 of unique BCR RNA sequence count. Usage is the proportion of all BCR RNA sequences falling into either a specific isotype or v-segment category (calculated separately for isotypes and v-segments). Class switching proportion is the proportion of all BCR RNA sequences that belong to IgA, IgG, or IgE isotypes and have evidence of somatic hypermutation (>1 mutation relative to the IMGT reference database). CDR3 length is the length of the CDR3 sequence in nucleotides. Q-value is calculated using the Benjamini-Hochberg method. |  |  |  |  |  |  |  |  |  |

Current CS also showed suggestive association to increased secretory IgA usage and decreased IgM, with several other borderline but non-significant associations. Vaping showed a suggestive association to decreased CDR3 length in IgE antibody transcripts as well as several borderline but non-significant associations (Figure E2). Overall, vaping and CS showed a similar trend in effect sizes compared to dual-use, suggesting that the effects were similar but less pronounced in current smokers and vapers relative to dual-users. When comparing dual-use to CS, we found no significant associations with BCR-measures. When comparing dual-use to vaping, we found dual-use was associated with lower IgE CDR3 length (β = −7.14 (SE: 2.28, adjusted p-value = 0.034)) and higher IgA2 usage (β = 0.032 (SE: 0.011, adj. p-value = 0.052)).

To examine the appropriateness of using former rather than never smokers as the reference group, we performed multivariable linear regressions comparing isotype usage in former versus never smoking individuals (Table E7), which demonstrated no significant differences between these groups after adjusting for multiple comparisons, suggesting that smoking effects on class switching may resolve after cessation.

### Associations between Sex and Self-Reported Race on B cell antibody production

In multivariable models, some of the strongest observed associations for BCR-seq measures were with self-reported race (Table 3). Comparing self-reported NHW versus AA participants, NHW-identifying participants had decreased usage of IgG1, IgG2, and IgG3 isotypes and increased usage of IgM (Figure 3). To investigate the extent to which these associations may be driven by variables related to income or socioeconomic status, we repeated the analysis adjusting for self-reported income level and area deprivation index, and 4 of the 6 significant associations remained significant, and all 6 associations had a consistent effect direction (Table E8). After adjusting for principal components of genetic ancestry, the isotype usage associations with race were attenuated, though notably, the principal component variables were also not associated with isotype usage. We observed no interaction between self-identified race and CS, vaping, or dual-use on isotype usage (all p > 0.05).

**Figure 3.**
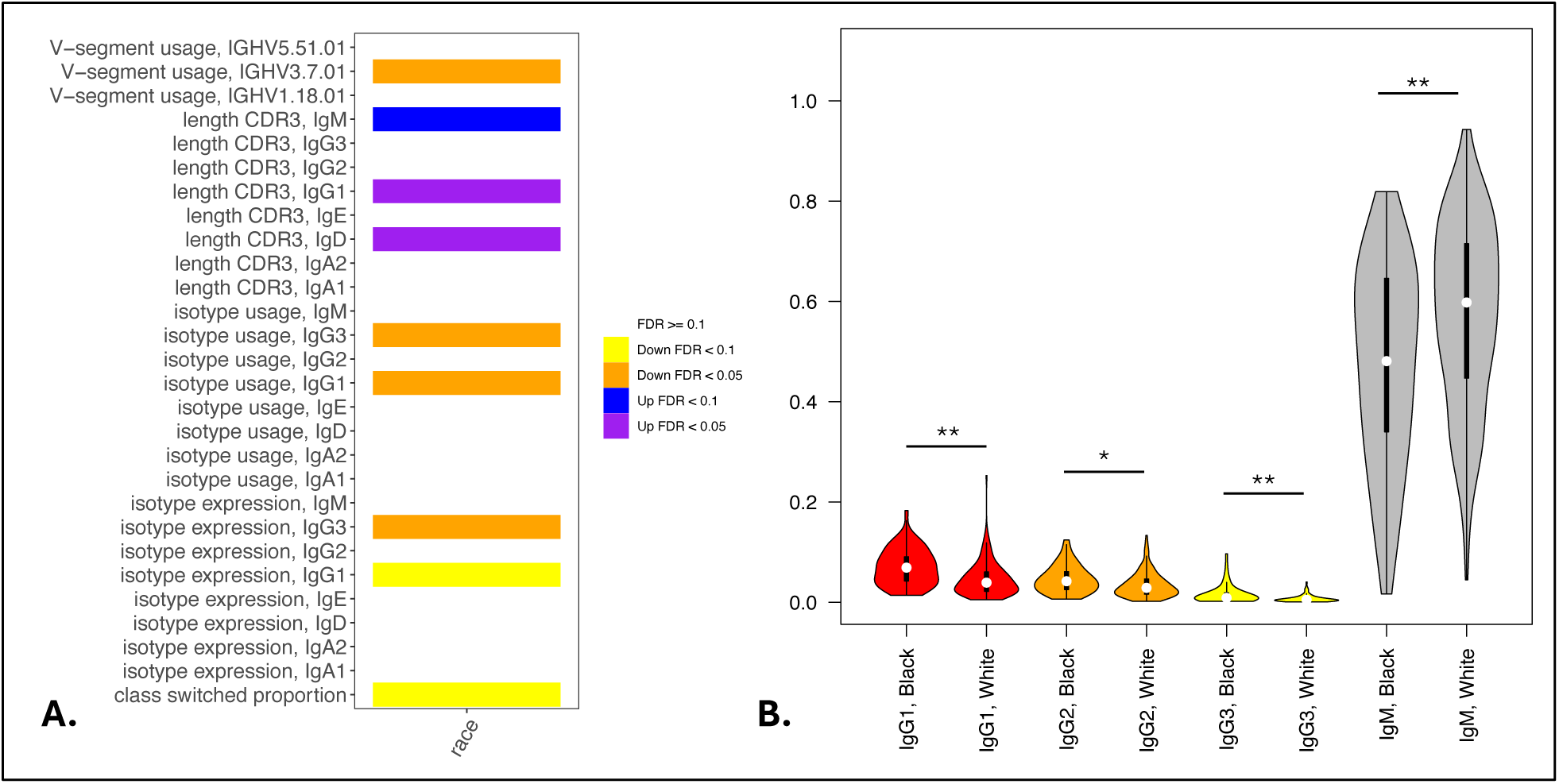
Associations of BCR Measures with Self-Reported Race. Significant associations between BCR measures and self-reported AA or NHW participants from multivariable models analyzing class switching, isotype expression and usage, V-segment usage, and CDR3 length (in nucleotides) are shown in Panel A. Panel B shows IgG and IgM isotype usage by self-reported race. Significance is assessed by t-tests. * p<=0.05, ** p<=0.005, ns p>0.05.

**Table 3.**
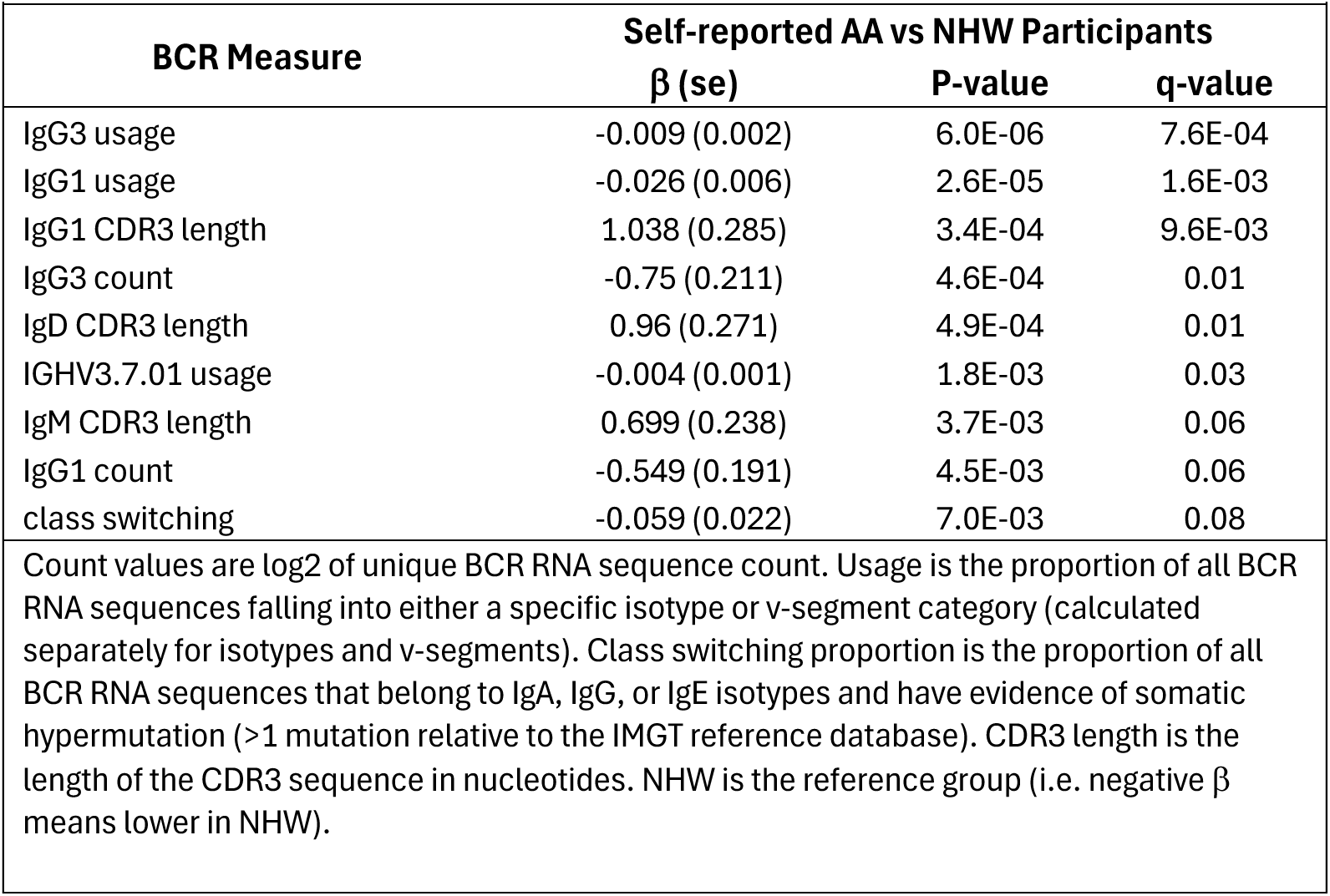
Significant BCR Associations to Self-Reported Race.

In the sex analysis, we observed one significant association in which male compared to female sex was significantly associated with decreased CDR3 length in IgM isotype sequences (β −0.63 (SE: 0.18, p=0.0008)).

### Associations to COPD and Related Phenotypes

We also examined associations between BCR-seq measures and age, COPD affection status, and CT-related measures of emphysema and airway wall thickness. We observed a significant univariate association between COPD and increased usage of the IGHV5.51.01 V-segment and suggestive associations with increased class switching and a shift from IgM to IgA (Table E9). Similar but non-significant trends were associated with the CT-quantified airway wall thickness (Table E10). However, these associations were not significant in models adjusting for CS/vaping behavior, age, sex, and race. No significant associations were observed with CT emphysema measures.

## Discussion

In this study of 234 individuals with B cell receptor sequencing (BCR-seq) and cigarette smoking (CS), vaping, and COPD-related outcome data, we observed significant effects of CS and vaping leading to increased IgA expression and usage, increased class switching, and lower antibody diversity indicating greater clonal activation of specific B cell populations. Taken together, these results demonstrate the potential dangers of dual-use compared to single product use, an under-recognized but important public health concern.

Our study demonstrates associations between dual-use and increased production of IgA, increased class switching, increased usage of specific V-segments and clonal expansion of B cells. These results point to increased B cell activation and increased production of secretory IgA (i.e. IgA2), consistent with mucosal exposure to compounds from vaping devices and combustible cigarettes. We note that CS was suggestively associated with increased IgA usage, and vaping also showed a borderline association with a consistent effect direction (adjusted p=0.12). It is possible that in a larger study these associations would reach statistical significance. IgA is secreted by the airway mucosa and is important in lung immune defense against pathogens^19^. Our data suggest that CS and vaping trigger similar host immune responses with a greater effect observed in participants engaged in dual-use. Indeed, many patients who use e-cigarettes for smoking cessation will smoke tobacco cigarettes and vape, and our results underscore the importance of understanding the health effects of dual-use specifically, as well as vaping and CS alone.

Our findings are consistent with previous research demonstrating that CS increases IgA production in blood and lung^20,21^. Higher levels of class-switched memory B cells have been observed in individuals who smoke compared to former and never smokers, irrespective of COPD status^22^. Vaping is associated with increased circulating club cell protein and decreased transcutaneous oxygen tension^3^, increased IL-10 and TNF-α^23^, and methylation changes that may cause long-term alterations in cytokine levels^24,25^. Vaping has also been associated with increased expression of 191 inflammatory proteins from bronchoalveolar lavage fluid, including MUC5AC, which is important in mucin production^26^. Clinically, vaping is associated with acute lung injury^2,27^, decreased FEV_1_/FVC and peak expiratory flow in asthmatics^23^, and chronic bronchitis^26^. However, the implications of dual-use on adaptive immunity are an important contribution of our study.

The role of adaptive immunity and B cells in COPD and emphysema pathogenesis is well-recognized^10,19,28,29^. Although we observed several univariable associations of blood B cell transcriptomics to COPD and related phenotypes, none of these associations remained significant in multivariable models after adjustment for multiple comparisons. While seemingly in contrast to the well-known increase in lung lymphoid follicles and B cell infiltration in severe COPD^10–12,28,29^, this lack of significant associations is not surprising, as by definition the presence of germinal centers in lung lymphoid follicles indicates local B cell division and maturation. Indeed, antigen exposure within the lung leads to local recruitment of memory (and perhaps naïve) B cells, local expansion of B cells and plasma cells within the airway tissue, and subsequent production of antigen-specific antibodies that tend to stay in the lung before getting into the bloodstream^10,30^. Since our analysis is limited to the transcriptome (cross-sectionally) of circulating B-cells, our results do not rule out spillover of antibody produced by lung-resident B-cells into the blood, and other COPD-specific inflammatory changes in blood such as neutrophilia and the increase in several RNA and protein biomarkers are well-documented ^31–33^. Our results suggest that the circulating B cell population in participants with COPD does not show large COPD-specific changes. Since COPD is strongly associated with CS and older age, participants with COPD would not be expected to have “normal” B cell function reflective of good health, but rather they would have B cell alterations that are characteristic of individuals with similar age and smoking history.

An intriguing aspect of BCR-seq is the ability to characterize the B cell response at the level of specific B cell clones. It is of interest that multiple associations between dual-use and race were observed with the usage of specific V-segments. For example, dual-use was strongly associated with increased usage of IGHV5.51.01 which has been associated with immune responses to parainfluenza viruses^34^, a common cause of upper respiratory illness that can cause severe respiratory illness in older or immunocompromised individuals.

Our finding of higher levels of IgG1 and IgG3 in AA compared to NHW participants agrees with a previous report^35^, which we extend by demonstrating this association in the context of smoking/vaping behaviors. Further, we observed no interaction between race and smoking/vaping variables on isotype usage, though it is important to note that our study is not well-powered to detect interactions of modest effect. Despite the lack of a large literature on racial differences in B cell function, this topic is of substantial interest due to the increased risk of multiple myeloma in AA individuals^36^. IgG1 and IgG3 both have excellent complement activation and opsonization capabilities, and IgG3 is a potent immune effector, suggesting differences in response to pathogens and toxins. As certain continuous traits, such as height and skin color, can vary with a person’s ancestral geographic origins, the observed associations with self-identified race could represent complex gene-by-environment interactions, or like all associations, could just be due to unmeasured confounding. In sensitivity analyses adjusting for income levels and area deprivation index, the associations with self-reported race and smoking/vaping status remained significant. We further adjusted for principal components of genetic ancestry, which attenuated race associations with isotype usage; however, there was no association of the principal components of genetic ancestry with isotype usage, suggesting that genetic principal components alone do not account for the observed differences. These findings need to be confirmed by future studies that focus on identifying the potential biological and socio-economic factors driving these differences as well as exploring forms of genetic analysis that account for admixture. Such studies would likely yield useful data for assessing disease risk and understanding vaccination response. To our knowledge, this is the largest study to date of BCR-seq in humans or model systems.

The strengths of this study include the novel use of BCR-seq in a large, deeply phenotyped cohort of participants engaged in current CS, vaping, or both. One limitation is that we did not have a suitable replication cohort, but our current findings highlight the need to obtain BCR-seq data longitudinally and in additional cohorts. We were not able to compare B cell activation in lung versus blood, which is important for understanding the role of adaptive immunity in CS, vaping, and COPD pathogenesis. Single-cell and spatial transcriptomic or proteomic data would provide greater resolution of the adaptive immune responses to CS and vaping. T cell receptor sequencing that coincides with BCR-seq would provide a more comprehensive view of adaptive immune responses in this context as well. A larger sample size would be desirable to examine COPD-related outcomes, particularly longitudinal outcomes such as FEV_1_ decline, mortality, and exacerbations.

In conclusion, we observed that CS and vaping each enhance B cell activation, and that dual-users show a trend towards greater effects than either alone. Self-identified race was strongly associated with IgG isotype usage. These findings highlight associations between B cell activation and antibody transcription, suggesting potential differences in immune and vaccine responses linked to CS, vaping, and self-identified race.

## Data Availability

All data are publicly available through dbGaP (COPDGene: phs000179.v1.p1).

