## Supplementary appendix for "B cell immune repertoire sequencing in tobacco cigarette smoking, vaping, and chronic obstructive pulmonary disease in the COPDGene cohort"


Funding and Acknowledgements 2
Supplementary Methods 4
B cell receptor repertoire sequencing (BCR-seq) library preparation 4
AIRR-seq data processing 5
Supplementary Results 6
Supplementary Figures 6
Figure E1 6
Figure E2 7
Supplementary Tables 8
Table E1 8
Table E2 11
Table E3 14
Table E4 16
Table E5 17
Table E6 20
Table E7 21
Table E8 23
Table E9 25
Table E10 26
References 27

### Funding and Acknowledgements

**COPDGene Phase 3 Grant Support and Disclaimer**

The COPDGene study (NCT00608764) is supported by grants from the NHLBI (U01HL089897 and U01HL089856) and by NIH contract 75N92023D00011. The content is solely the responsibility of the authors and does not necessarily represent the official views of the National Heart, Lung, and Blood Institute or the National Institutes of Health.

**COPD Foundation Funding**

COPDGene is also supported by the COPD Foundation through contributions made to an Industry Advisory Board that has included AstraZeneca, Bayer Pharmaceuticals, Boehringer-Ingelheim, Genentech, GlaxoSmithKline, Novartis, Pfizer, and Sunovion.

**COPDGene® Investigators – Core Units**

*Administrative Center*: James D. Crapo, MD (PI); Edwin K. Silverman, MD, PhD (PI); Barry J. Make, MD; Elizabeth A. Regan, MD, PhD

*Genetic Analysis Center*: Terri H. Beaty, PhD; Peter J. Castaldi, MD, MSc; Michael H. Cho, MD, MPH; Dawn L. DeMeo, MD, MPH; Adel El Boueiz, MD, MMSc; Marilyn G. Foreman, MD, MS; Auyon Ghosh, MD; Lystra P. Hayden, MD, MMSc; Craig P. Hersh, MD, MPH; Jacqueline Hetmanski, MS; Brian D. Hobbs, MD, MMSc; John E. Hokanson, MPH, PhD; Wonji Kim, PhD; Nan Laird, PhD; Christoph Lange, PhD; Sharon M. Lutz, PhD; Merry-Lynn McDonald, PhD; Dmitry Prokopenko, PhD; Matthew Moll, MD, MPH; Jarrett Morrow, PhD; Dandi Qiao, PhD; Elizabeth A. Regan, MD, PhD; Aabida Saferali, PhD; Phuwanat Sakornsakolpat, MD; Edwin K. Silverman, MD, PhD; Emily S. Wan, MD; Jeong Yun, MD, MPH

*Imaging Center*: Juan Pablo Centeno; Jean-Paul Charbonnier, PhD; Harvey O. Coxson, PhD; Craig J. Galban, PhD; MeiLan K. Han, MD, MS; Eric A. Hoffman, Stephen Humphries, PhD; Francine L. Jacobson, MD, MPH; Philip F. Judy, PhD; Ella A. Kazerooni, MD; Alex Kluiber; David A. Lynch, MB; Pietro Nardelli, PhD; John D. Newell, Jr., MD; Aleena Notary; Andrea Oh, MD; Elizabeth A. Regan, MD, PhD; James C. Ross, PhD; Raul San Jose Estepar, PhD; Joyce Schroeder, MD; Jered Sieren; Berend C. Stoel, PhD; Juerg Tschirren, PhD; Edwin Van Beek, MD, PhD; Bram van Ginneken, PhD; Eva van Rikxoort, PhD; Gonzalo Vegas Sanchez- Ferrero, PhD; Lucas Veitel; George R. Washko, MD; Carla G. Wilson, MS;

*PFT QA Center, Salt Lake City, UT*: Robert Jensen, PhD

*Data Coordinating Center and Biostatistics*, *National Jewish Health, Denver, CO*: Douglas Everett, PhD; Jim Crooks, PhD; Katherine Pratte, PhD; Matt Strand, PhD; Carla G. Wilson, MS

*Epidemiology Core*, *University of Colorado Anschutz Medical Campus, Aurora, CO*: John E. Hokanson, MPH, PhD; Erin Austin, PhD; Gregory Kinney, MPH, PhD; Sharon M. Lutz, PhD; Kendra A. Young, PhD

*Mortality Adjudication Core:* Surya P. Bhatt, MD; Jessica Bon, MD; Alejandro A. Diaz, MD, MPH; MeiLan K. Han, MD, MS; Barry Make, MD; Susan Murray, ScD; Elizabeth Regan, MD; Xavier Soler, MD; Carla G. Wilson, MS

*Biomarker Core*: Russell P. Bowler, MD, PhD; Katerina Kechris, PhD; Farnoush Banaei-Kashani, PhD

**COPDGene® Investigators – Clinical Centers**

*Ann Arbor VA:* Jeffrey L. Curtis, MD; Perry G. Pernicano, MD

*Baylor College of Medicine, Houston, TX*: Nicola Hanania, MD, MS; Mustafa Atik, MD; Aladin Boriek, PhD; Kalpatha Guntupalli, MD; Elizabeth Guy, MD; Amit Parulekar, MD;

*Brigham and Women's Hospital, Boston, MA*: Dawn L. DeMeo, MD, MPH; Craig Hersh, MD, MPH; Francine L. Jacobson, MD, MPH; George Washko, MD

*Columbia University, New York, NY*: R. Graham Barr, MD, DrPH; John Austin, MD; Belinda D'Souza, MD; Byron Thomashow, MD

*Duke University Medical Center, Durham, NC*: Neil MacIntyre, Jr., MD; H. Page McAdams, MD; Lacey Washington, MD

*HealthPartners Research Institute, Minneapolis, MN*: Charlene McEvoy, MD, MPH; Joseph Tashjian, MD

*Johns Hopkins University, Baltimore, MD*: Robert Wise, MD; Robert Brown, MD; Nadia N. Hansel, MD, MPH; Karen Horton, MD; Allison Lambert, MD, MHS; Nirupama Putcha, MD, MHS

*Lundquist Institute for Biomedical Innovation at Harbor UCLA Medical Center, Torrance, CA*: Richard Casaburi, PhD, MD; Alessandra Adami, PhD; Matthew Budoff, MD; Hans Fischer, MD; Janos Porszasz, MD, PhD; Harry Rossiter, PhD; William Stringer, MD
*Michael E. DeBakey VAMC, Houston, TX*: Amir Sharafkhaneh, MD, PhD; Charlie Lan, DO
*Minneapolis VA:* Christine Wendt, MD; Brian Bell, MD; Ken M. Kunisaki, MD, MS

*Morehouse School of Medicine, Atlanta, GA*: Eric L. Flenaugh, MD; Hirut Gebrekristos, PhD; Mario Ponce, MD; Silanath Terpenning, MD; Gloria Westney, MD, MS

*National Jewish Health, Denver, CO*: Russell Bowler, MD, PhD; David A. Lynch, MB *Reliant Medical Group, Worcester, MA*: Richard Rosiello, MD; David Pace, MD

*Temple University, Philadelphia, PA:* Gerard Criner, MD; David Ciccolella, MD; Francis Cordova, MD; Chandra Dass, MD; Gilbert D'Alonzo, DO; Parag Desai, MD; Michael Jacobs, PharmD; Steven Kelsen, MD, PhD; Victor Kim, MD; A. James Mamary, MD; Nathaniel Marchetti, DO; Aditi Satti, MD; Kartik Shenoy, MD; Robert M. Steiner, MD; Alex Swift, MD; Irene Swift, MD; Maria Elena Vega-Sanchez, MD

*University of Alabama, Birmingham, AL:* Mark Dransfield, MD; William Bailey, MD; Surya P. Bhatt, MD; Anand Iyer, MD; Hrudaya Nath, MD; J. Michael Wells, MD

*University of California, San Diego, CA*: Douglas Conrad, MD; Xavier Soler, MD, PhD; Andrew Yen, MD

*University of Iowa, Iowa City, IA*: Alejandro P. Comellas, MD; Karin F. Hoth, PhD; John Newell, Jr., MD; Brad Thompson, MD

*University of Michigan, Ann Arbor, MI:* MeiLan K. Han, MD MS; Ella Kazerooni, MD MS; Wassim Labaki, MD MS; Craig Galban, PhD; Dharshan Vummidi, MD

*University of Minnesota, Minneapolis, MN*: Joanne Billings, MD; Abbie Begnaud, MD; Tadashi Allen, MD

*University of Pittsburgh, Pittsburgh, PA*: Frank Sciurba, MD; Jessica Bon, MD; Divay Chandra, MD, MSc; Joel Weissfeld, MD, MPH

*University of Texas Health, San Antonio, San Antonio, TX*: Antonio Anzueto, MD; Sandra Adams, MD; Diego Maselli-Caceres, MD; Mario E. Ruiz, MD; Harjinder Singh

### Supplementary Methods

#### B cell receptor repertoire sequencing (BCR-seq) library preparation

Library preparation was performed according to Vollmers et al.[1]. Briefly, a pooled set of 5 isotype-specific IGH constant region primers (RT_pool), containing 12 random nucelotides (nt) and partial Illumina adapters were added to the RNA samples, incubated at 72°C for 3 min then immediately placed on ice for 2 min. First-strand cDNA synthesis was then performed using Smartscribe reverse transcriptase (Takara) according to manufacturer's instructions. Second-strand cDNA synthesis was done using Phusion HiFi DNA polymerase (ThermoFisher) using a pool of six IGH variable region primers (V_FR1_pool) and a pool of five IGH constant region primers (C_pool), both containing 12 random nts and partial Illumina adapters. The reaction was incubated using the following heat cycle program: [98°C for 4 min, 52°C for 1 min, 72°C for

5 min]x2. The resulting double-stranded cDNA was purified and size-selected for molecules > 350bp using Zymo Select-a-Size columns. Finally, the double-stranded cDNA was amplified using 2x kapa hifi hotstart readymix (Roche) using primers containing complete Illumina adapters indexes (NexteraA_Index and NexteraB_Index primers) and the following heat cycle program: 95C for 3 mins, [98C for 20s, 67C for 15s, 72C for 1min]x26, 72C for 5 mins. PCR products were purified once using Ampure XP beads at a 0.7:1 ratio then pooled for multiplexed sequencing using the Illumina MiSeq sequencer and 2x300 run kits and flowcells.

### AIRR-seq data processing

We downloaded from IMGT the reference germline sequences for IGH V (IMGT-gapped), D and J genes and created blast database files following instructions from NCBI IgBLAST. Blast database for IGH C genes were directly downloaded from NCBI IgBLAST release ftp site. Fasta files of preprocessed AIRR-seq reads were then aligned to IGH V, D, J and C genes using igblastn command line program from IgBLAST (version 1.19.0). We further filtered aligned reads keeping only those with non-missing v_support and j_support fields, v_support >= 1e-50 and predicted to be productive. We used the spectralClones function from the scoper R package to infer clonal relationships between the sequence reads.

### Supplementary Results

### Supplementary Figures

*Figure E1*: Hill biodiversity numbers show less antibody diversity (and more clonal expansion) in dual users relative to other smoking/vaping groups. The top panel is a boxplot showing that the dual users (i.e., vaping + smoking) have lower log-Hill values than other groups. In the bottom panel, we collapsed all other groups and compared the hill numbers in a continuous fashion over a range of q-values.

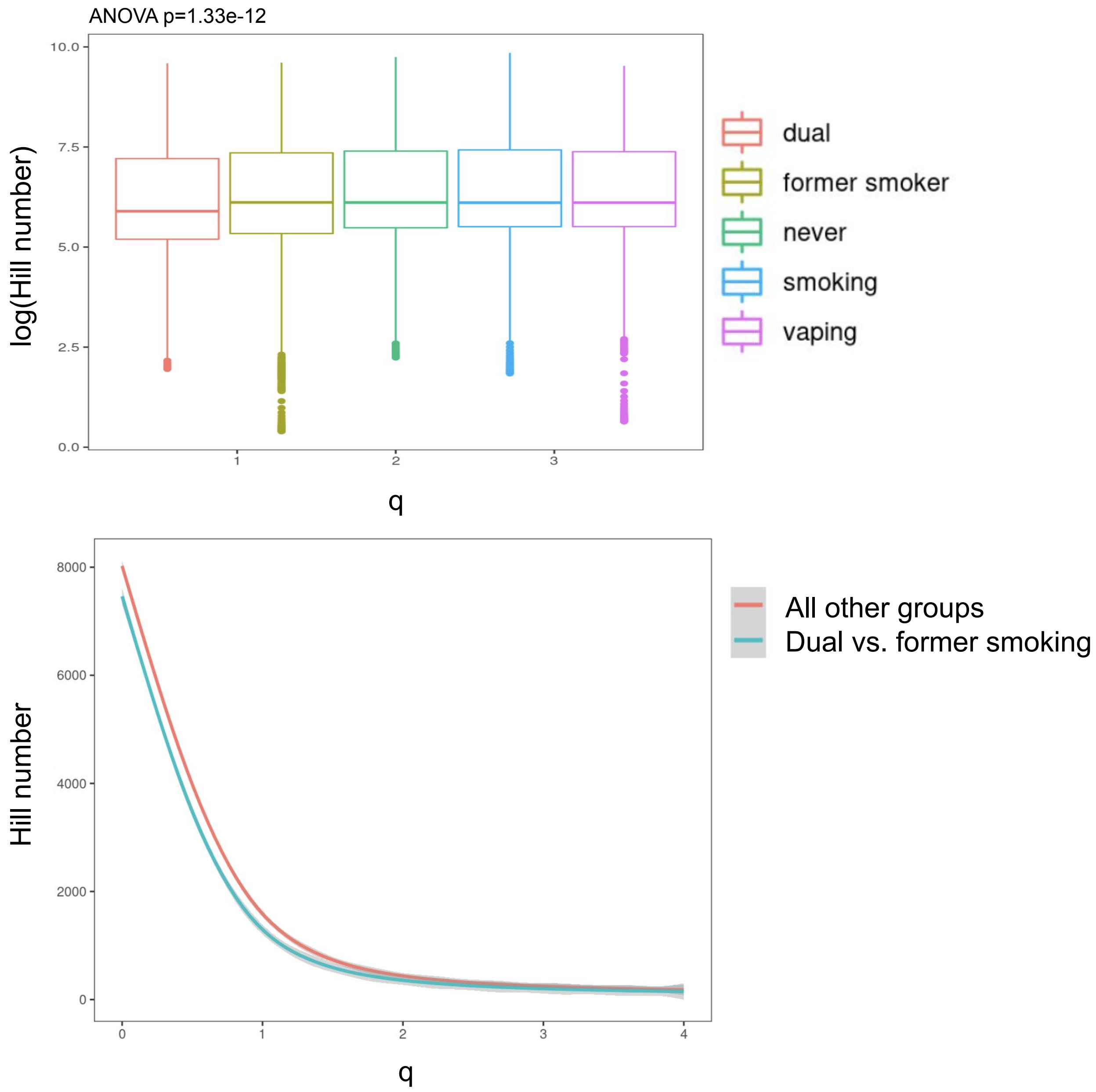

*Figure E2*: Difference in CDR3 Length between current vaping and former smoking participants

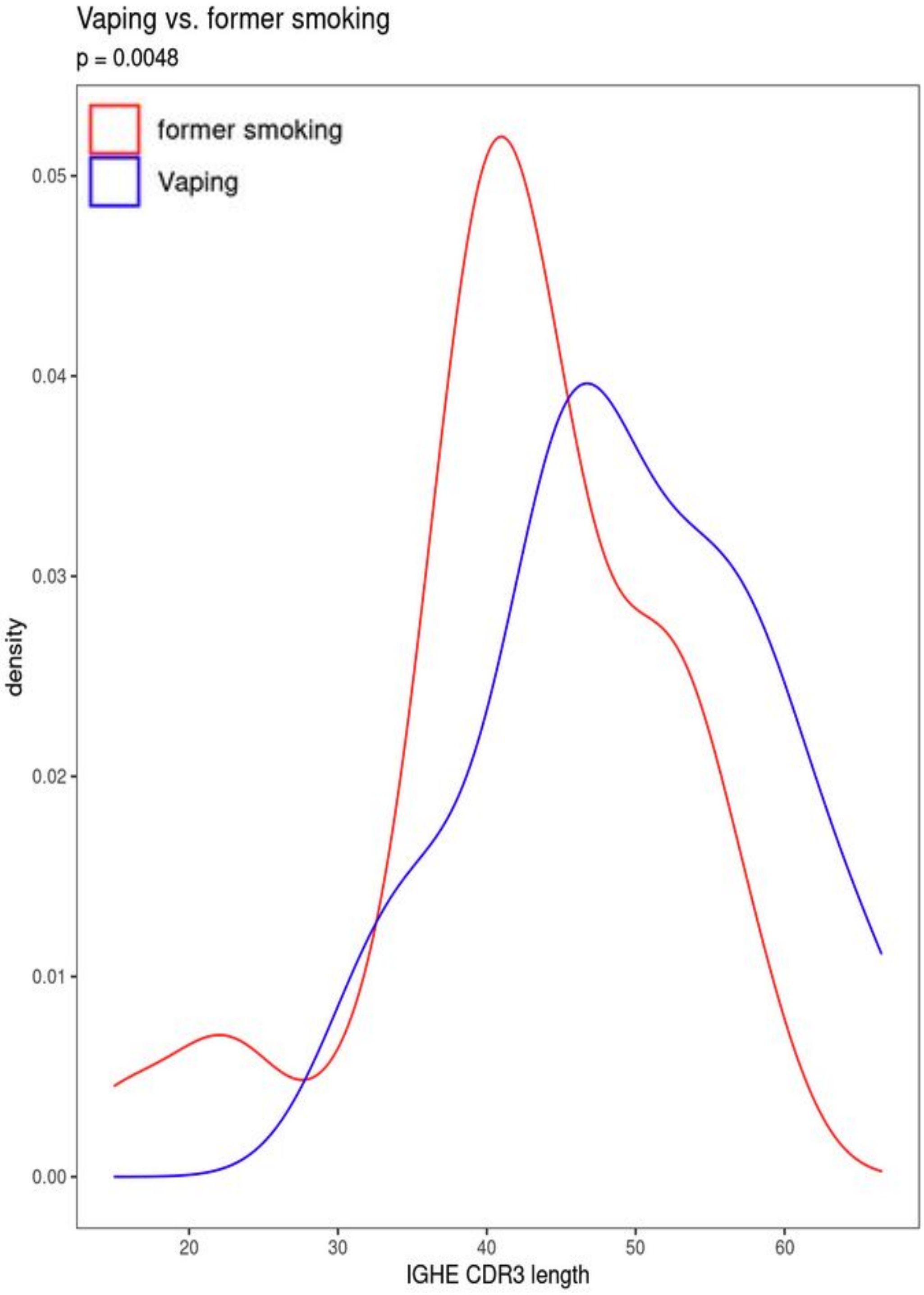

### Supplementary Tables

*Table E1*. **Regression Models for Isotype Usage with Independent Variables of Interest**

| Dependent Variable | Independent Variables | beta (se) | P-value | q-value |
|---|---|---|---|---|
| IGHA1 usage | age | 0 (0.001) | 9.4E-01 | 9.7E-01 |
| | sex | 0.019 (0.015) | 2.1E-01 | 5.3E-01 |
| | race | -0.023 (0.019) | 2.2E-01 | 5.5E-01 |
| | dual use | 0.095 (0.023) | 7.5E-05 | 2.7E-03 |
| | smoking | 0.061 (0.025) | 1.4E-02 | 1.1E-01 |
| | vaping | 0.051 (0.024) | 3.5E-02 | 1.9E-01 |
| | GOLD 1 vs 0 | 0.031 (0.028) | 2.6E-01 | 6.1E-01 |
| | GOLD 2-4 vs 0 | 0.033 (0.018) | 7.5E-02 | 3.1E-01 |
| | GOLD PrISM vs 0 | -0.019 (0.03) | 5.2E-01 | 7.9E-01 |
| IGHA2 usage | age | 0 (0) | 9.2E-01 | 9.7E-01 |
| | sex | 0.018 (0.007) | 1.0E-02 | 1.1E-01 |
| | race | 0 (0.009) | 9.9E-01 | 9.9E-01 |
| | dual use | 0.059 (0.011) | 2.8E-07 | 7.0E-05 |
| | smoking | 0.035 (0.012) | 3.3E-03 | 5.2E-02 |
| | vaping | 0.027 (0.012) | 1.9E-02 | 1.2E-01 |
| | GOLD 1 vs 0 | 0.013 (0.013) | 3.3E-01 | 6.6E-01 |
| | GOLD 2-4 vs 0 | 0.016 (0.009) | 7.3E-02 | 3.1E-01 |
| | GOLD PrISM vs 0 | -0.016 (0.014) | 2.6E-01 | 6.1E-01 |
| IGHD usage | age | 0 (0.001) | 8.9E-01 | 9.7E-01 |
| | sex | -0.001 (0.007) | 9.3E-01 | 9.7E-01 |
| | race | 0.004 (0.01) | 6.9E-01 | 8.9E-01 |
| | dual use | -0.015 (0.012) | 1.9E-01 | 5.2E-01 |
| | smoking | 0 (0.012) | 9.8E-01 | 9.9E-01 |
| | vaping | -0.002 (0.012) | 8.4E-01 | 9.6E-01 |

| | | | | |
|---|---|---|---|---|
| | GOLD 1 vs 0 | 0 (0.014) | 9.9E-01 | 9.9E-01 |
| | GOLD 2-4 vs 0 | -0.007 (0.009) | 4.4E-01 | 7.5E-01 |
| | GOLD PrISM vs 0 | 0.009 (0.015) | 5.5E-01 | 7.9E-01 |
| IGHE usage | age | 0 (0) | 8.2E-02 | 3.1E-01 |
| | sex | 0 (0) | 8.7E-02 | 3.2E-01 |
| | race | 0 (0) | 1.3E-02 | 1.1E-01 |
| | dual use | 0 (0) | 2.7E-01 | 6.1E-01 |
| | smoking | 0 (0) | 9.7E-01 | 9.9E-01 |
| | vaping | 0 (0) | 8.6E-01 | 9.6E-01 |
| | GOLD 1 vs 0 | 0 (0) | 5.7E-01 | 8.0E-01 |
| | GOLD 2-4 vs 0 | 0 (0) | 3.4E-01 | 6.7E-01 |
| | GOLD PrISM vs 0 | 0 (0) | 6.6E-01 | 8.7E-01 |
| IGHG1 usage | age | 0 (0) | 1.6E-01 | 4.8E-01 |
| | sex | 0.003 (0.005) | 5.4E-01 | 7.9E-01 |
| | race | -0.026 (0.006) | 2.6E-05 | 1.6E-03 |
| | dual use | 0.016 (0.007) | 3.2E-02 | 1.7E-01 |
| | smoking | 0.011 (0.008) | 1.6E-01 | 4.8E-01 |
| | vaping | 0.018 (0.008) | 2.0E-02 | 1.3E-01 |
| | GOLD 1 vs 0 | -0.008 (0.009) | 3.6E-01 | 6.7E-01 |
| | GOLD 2-4 vs 0 | 0.005 (0.006) | 4.0E-01 | 7.2E-01 |
| | GOLD PrISM vs 0 | -0.009 (0.01) | 3.4E-01 | 6.7E-01 |
| IGHG2 usage | age | 0 (0) | 5.5E-01 | 7.9E-01 |
| | sex | 0.003 (0.003) | 3.9E-01 | 7.1E-01 |
| | race | -0.01 (0.004) | 2.4E-02 | 1.4E-01 |
| | dual use | 0.013 (0.006) | 1.6E-02 | 1.2E-01 |
| | smoking | 0.004 (0.006) | 5.3E-01 | 7.9E-01 |
| | vaping | 0.003 (0.006) | 5.5E-01 | 7.9E-01 |
| | GOLD 1 vs 0 | -0.002 (0.007) | 7.4E-01 | 9.1E-01 |

| | | | | |
|---|---|---|---|---|
| | GOLD 2-4 vs 0 | 0.005 (0.004) | 2.9E-01 | 6.3E-01 |
| | GOLD PrISM vs 0 | 0.001 (0.007) | 8.4E-01 | 9.6E-01 |
| IGHG3 usage | age | 0 (0) | 8.4E-01 | 9.6E-01 |
| | sex | 0.001 (0.002) | 4.1E-01 | 7.2E-01 |
| | race | -0.009 (0.002) | 6.0E-06 | 7.6E-04 |
| | dual use | 0.002 (0.002) | 5.3E-01 | 7.9E-01 |
| | smoking | 0.003 (0.003) | 2.1E-01 | 5.3E-01 |
| | vaping | 0.002 (0.002) | 3.6E-01 | 6.8E-01 |
| | GOLD 1 vs 0 | 0.005 (0.003) | 7.9E-02 | 3.1E-01 |
| | GOLD 2-4 vs 0 | 0.004 (0.002) | 5.6E-02 | 2.5E-01 |
| | GOLD PrISM vs 0 | -0.001 (0.003) | 8.5E-01 | 9.6E-01 |
| IGHM usage | age | -0.001 (0.002) | 4.7E-01 | 7.7E-01 |
| | sex | -0.05 (0.024) | 4.1E-02 | 2.1E-01 |
| | race | 0.074 (0.031) | 1.8E-02 | 1.2E-01 |
| | dual use | -0.173 (0.038) | 1.1E-05 | 8.9E-04 |
| | smoking | -0.113 (0.04) | 5.5E-03 | 7.3E-02 |
| | vaping | -0.087 (0.04) | 2.9E-02 | 1.7E-01 |
| | GOLD 1 vs 0 | -0.024 (0.045) | 5.9E-01 | 8.1E-01 |
| | GOLD 2-4 vs 0 | -0.047 (0.03) | 1.2E-01 | 4.0E-01 |
| | GOLD PrISM vs 0 | 0.024 (0.049) | 6.2E-01 | 8.4E-01 |

Usage is the proportion of all BCR RNA sequences falling into either a specific isotype or v-segment category (calculated separately for isotypes and v-segments). Sex coded as (0=female, 1=male). Race coded as (0=self reported Black race, 1=self reported White race). PrISM - Preserved ratio impaired spirometry.

*Table E2*. **Regression Models for Isotype Expression with Independent Variabes of Interest**

| Dependent Variable | Independent Variables | beta (se) | P-value | q-value |
|---|---|---|---|---|
| IGHA1 expression | age | -0.016 (0.012) | 1.8E-01 | 5.2E-01 |
| | sex | 0.075 (0.177) | 6.7E-01 | 8.8E-01 |
| | race | -0.07 (0.229) | 7.6E-01 | 9.2E-01 |
| | dual use | 0.724 (0.281) | 1.1E-02 | 1.1E-01 |
| | smoking | 0.59 (0.294) | 4.6E-02 | 2.4E-01 |
| | vaping | 0.167 (0.291) | 5.7E-01 | 8.0E-01 |
| | GOLD 1 vs 0 | -0.189 (0.339) | 5.8E-01 | 8.0E-01 |
| | GOLD 2-4 vs 0 | 0.018 (0.218) | 9.3E-01 | 9.7E-01 |
| | GOLD PrISM vs 0 | -0.508 (0.359) | 1.6E-01 | 4.8E-01 |
| IGHA2 expression | age | -0.012 (0.013) | 3.4E-01 | 6.7E-01 |
| | sex | 0.18 (0.185) | 3.3E-01 | 6.6E-01 |
| | race | 0.033 (0.242) | 8.9E-01 | 9.7E-01 |
| | dual use | 0.879 (0.294) | 3.1E-03 | 5.2E-02 |
| | smoking | 0.766 (0.306) | 1.3E-02 | 1.1E-01 |
| | vaping | 0.432 (0.301) | 1.5E-01 | 4.8E-01 |
| | GOLD 1 vs 0 | -0.005 (0.346) | 9.9E-01 | 9.9E-01 |
| | GOLD 2-4 vs 0 | 0.048 (0.229) | 8.4E-01 | 9.6E-01 |
| | GOLD PrISM vs 0 | -0.746 (0.373) | 4.7E-02 | 2.4E-01 |
| IGHD expression | age | -0.019 (0.01) | 5.3E-02 | 2.4E-01 |
| | sex | -0.149 (0.142) | 2.9E-01 | 6.3E-01 |
| | race | 0.147 (0.183) | 4.2E-01 | 7.4E-01 |
| | dual use | -0.273 (0.226) | 2.3E-01 | 5.6E-01 |
| | smoking | 0.099 (0.236) | 6.8E-01 | 8.8E-01 |
| | vaping | -0.253 (0.233) | 2.8E-01 | 6.2E-01 |
| | GOLD 1 vs 0 | 0.2 (0.267) | 4.5E-01 | 7.6E-01 |

| | | | | |
|---|---|---|---|---|
| | GOLD 2-4 vs 0 | -0.275 (0.176) | 1.2E-01 | 4.0E-01 |
| | GOLD PrISM vs 0 | -0.026 (0.288) | 9.3E-01 | 9.7E-01 |
| IGHE expression | age | -0.006 (0.015) | 6.8E-01 | 8.8E-01 |
| | sex | -0.005 (0.225) | 9.8E-01 | 9.9E-01 |
| | race | -0.203 (0.295) | 4.9E-01 | 7.9E-01 |
| | dual use | 0.492 (0.36) | 1.7E-01 | 5.1E-01 |
| | smoking | 0.595 (0.381) | 1.2E-01 | 4.0E-01 |
| | vaping | 0.379 (0.38) | 3.2E-01 | 6.5E-01 |
| | GOLD 1 vs 0 | -0.256 (0.437) | 5.6E-01 | 7.9E-01 |
| | GOLD 2-4 vs 0 | -0.288 (0.284) | 3.1E-01 | 6.4E-01 |
| | GOLD PrISM vs 0 | -0.824 (0.478) | 8.7E-02 | 3.2E-01 |
| IGHG1 expression | age | -0.01 (0.01) | 3.1E-01 | 6.4E-01 |
| | sex | -0.082 (0.149) | 5.8E-01 | 8.1E-01 |
| | race | -0.549 (0.191) | 4.5E-03 | 6.3E-02 |
| | dual use | 0.418 (0.237) | 7.9E-02 | 3.1E-01 |
| | smoking | 0.465 (0.248) | 6.3E-02 | 2.7E-01 |
| | vaping | 0.109 (0.246) | 6.6E-01 | 8.7E-01 |
| | GOLD 1 vs 0 | 0.134 (0.279) | 6.3E-01 | 8.5E-01 |
| | GOLD 2-4 vs 0 | -0.061 (0.183) | 7.4E-01 | 9.1E-01 |
| | GOLD PrISM vs 0 | -0.32 (0.31) | 3.0E-01 | 6.3E-01 |
| IGHG2 expression | age | -0.018 (0.011) | 1.0E-01 | 3.7E-01 |
| | sex | 0.058 (0.16) | 7.2E-01 | 9.0E-01 |
| | race | -0.293 (0.207) | 1.6E-01 | 4.8E-01 |
| | dual use | 0.357 (0.254) | 1.6E-01 | 4.8E-01 |
| | smoking | 0.275 (0.266) | 3.0E-01 | 6.3E-01 |
| | vaping | -0.004 (0.262) | 9.9E-01 | 9.9E-01 |
| | GOLD 1 vs 0 | 0.176 (0.3) | 5.6E-01 | 7.9E-01 |

| | | | | |
|---|---|---|---|---|
| | GOLD 2-4 vs 0 | -0.018 (0.198) | 9.3E-01 | 9.7E-01 |
| | GOLD PrISM vs 0 | -0.03 (0.324) | 9.3E-01 | 9.7E-01 |
| IGHG3 expression | age | -0.014 (0.011) | 2.0E-01 | 5.3E-01 |
| | sex | -0.127 (0.164) | 4.4E-01 | 7.5E-01 |
| | race | -0.75 (0.211) | 4.6E-04 | 1.0E-02 |
| | dual use | 0.331 (0.261) | 2.1E-01 | 5.3E-01 |
| | smoking | 0.368 (0.272) | 1.8E-01 | 5.1E-01 |
| | vaping | 0.072 (0.269) | 7.9E-01 | 9.4E-01 |
| | GOLD 1 vs 0 | 0.406 (0.307) | 1.9E-01 | 5.2E-01 |
| | GOLD 2-4 vs 0 | 0.199 (0.203) | 3.3E-01 | 6.6E-01 |
| | GOLD PrISM vs 0 | -0.199 (0.331) | 5.5E-01 | 7.9E-01 |
| IGHM expression | age | -0.022 (0.009) | 1.7E-02 | 1.2E-01 |
| | sex | -0.266 (0.136) | 5.1E-02 | 2.4E-01 |
| | race | 0.219 (0.175) | 2.1E-01 | 5.3E-01 |
| | dual use | -0.49 (0.216) | 2.4E-02 | 1.4E-01 |
| | smoking | -0.148 (0.226) | 5.1E-01 | 7.9E-01 |
| | vaping | -0.434 (0.223) | 5.2E-02 | 2.4E-01 |
| | GOLD 1 vs 0 | 0.112 (0.255) | 6.6E-01 | 8.7E-01 |
| | GOLD 2-4 vs 0 | -0.294 (0.168) | 8.1E-02 | 3.1E-01 |
| | GOLD PrISM vs 0 | -0.076 (0.276) | 7.8E-01 | 9.4E-01 |

Count values are log2 of unique BCR RNA sequence count. Sex coded as (0=female, 1=male). Race coded as (0=self reported Black race, 1=self reported White race). PrISM - Preserved ratio impaired spirometry.

*Table E3*. **Regression Models for V-segment Usage with Independent Variabes of Interest**

| Dependent Variable | Independent Variables | beta (se) | P-value | q-value |
|---|---|---|---|---|
| IGHV1.18.01 usage | age | 0 (0) | 5.1E-01 | 7.9E-01 |
| | sex | 0.001 (0.001) | 2.8E-01 | 6.2E-01 |
| | race | -0.003 (0.001) | 2.0E-02 | 1.3E-01 |
| | dual use | 0.005 (0.001) | 4.9E-04 | 1.0E-02 |
| | smoking | 0.004 (0.001) | 9.3E-03 | 1.1E-01 |
| | vaping | 0.004 (0.001) | 1.6E-02 | 1.2E-01 |
| | GOLD 1 vs 0 | 0.001 (0.002) | 3.7E-01 | 7.0E-01 |
| | GOLD 2-4 vs 0 | 0.001 (0.001) | 2.4E-01 | 5.8E-01 |
| | GOLD PrISM vs 0 | -0.001 (0.002) | 5.0E-01 | 7.9E-01 |
| IGHV3.7.01 usage | age | 0 (0) | 1.1E-01 | 3.8E-01 |
| | sex | 0.003 (0.001) | 1.3E-02 | 1.1E-01 |
| | race | -0.004 (0.001) | 1.8E-03 | 3.3E-02 |
| | dual use | 0.006 (0.002) | 3.2E-04 | 9.6E-03 |
| | smoking | 0.003 (0.002) | 5.3E-02 | 2.4E-01 |
| | vaping | 0.002 (0.002) | 2.8E-01 | 6.2E-01 |
| | GOLD 1 vs 0 | 0 (0.002) | 8.7E-01 | 9.6E-01 |
| | GOLD 2-4 vs 0 | -0.001 (0.001) | 5.5E-01 | 7.9E-01 |
| | GOLD PrISM vs 0 | 0 (0.002) | 9.2E-01 | 9.7E-01 |
| IGHV5.51.01 usage | age | 0 (0) | 3.5E-01 | 6.7E-01 |
| | sex | 0.003 (0.001) | 1.5E-02 | 1.2E-01 |
| | race | -0.001 (0.002) | 6.8E-01 | 8.8E-01 |
| | dual use | 0.009 (0.002) | 6.2E-05 | 2.6E-03 |
| | smoking | 0.005 (0.002) | 1.8E-02 | 1.2E-01 |
| | vaping | 0.006 (0.002) | 1.3E-02 | 1.1E-01 |
| | GOLD 1 vs 0 | -0.002 (0.003) | 4.6E-01 | 7.6E-01 |

| | | | | |
|---|---|---|---|---|
| | GOLD 2-4 vs 0 | 0.004 (0.002) | 3.0E-02 | 1.7E-01 |
| | GOLD PrISM vs 0 | -0.002 (0.003) | 5.5E-01 | 7.9E-01 |

Usage is the proportion of all BCR RNA sequences falling into either a specific isotype or v-segment category (calculated separately for isotypes and v-segments). Sex coded as (0=female, 1=male). Race coded as (0=self reported Black race, 1=self reported White race). PrISM - Preserved ratio impaired spirometry. V-segments where 75th percentile of usage is >0.01 were analyzed.

*Table E4*. **Regression Model for Class Switching with Independent Variabes of Interest**

| Dependent Variable | Independent Variables | beta (se) | P-value | q-value |
|---|---|---|---|---|
| class switching | age | 0.001 (0.001) | 4.7E-01 | 7.7E-01 |
| | sex | 0.033 (0.017) | 5.0E-02 | 2.4E-01 |
| | race | -0.059 (0.022) | 7.0E-03 | 8.4E-02 |
| | dual use | 0.111 (0.027) | 5.4E-05 | 2.6E-03 |
| | smoking | 0.064 (0.028) | 2.4E-02 | 1.4E-01 |
| | vaping | 0.05 (0.028) | 7.4E-02 | 3.1E-01 |
| | GOLD 1 vs 0 | 0.011 (0.032) | 7.2E-01 | 9.0E-01 |
| | GOLD 2-4 vs 0 | 0.028 (0.021) | 1.8E-01 | 5.2E-01 |
| | GOLD PrISM vs 0 | -0.013 (0.034) | 7.1E-01 | 8.9E-01 |

Class switching proportion is the proportion of all BCR RNA sequences that belong to IgA, IgG, or IgE isotypes and have evidence of somatic hypermutation (>1 mutation relative to the IMGT reference database). Sex coded as (0=female, 1=male). Race coded as (0=self reported Black race, 1=self reported White race). PrISM - Preserved ratio impaired spirometry.

*Table E5*. **Regression Models for CDR3 Length with Independent Variabes of Interest**

| Dependent Variable | Independent Variables | beta (se) | P-value | q-value |
|---|---|---|---|---|
| IGHA1 CDR3 Length | age | -0.008 (0.013) | 5.2E-01 | 7.9E-01 |
| | sex | -0.07 (0.185) | 7.0E-01 | 8.9E-01 |
| | race | 0.416 (0.238) | 8.2E-02 | 3.1E-01 |
| | dual use | 0.05 (0.295) | 8.7E-01 | 9.6E-01 |
| | smoking | -0.071 (0.309) | 8.2E-01 | 9.6E-01 |
| | vaping | 0.037 (0.305) | 9.0E-01 | 9.7E-01 |
| | GOLD 1 vs 0 | 0.134 (0.347) | 7.0E-01 | 8.9E-01 |
| | GOLD 2-4 vs 0 | 0.068 (0.228) | 7.7E-01 | 9.3E-01 |
| | GOLD PrISM vs 0 | 0.613 (0.385) | 1.1E-01 | 3.9E-01 |
| IGHA2 CDR3 Length | age | -0.013 (0.015) | 3.9E-01 | 7.1E-01 |
| | sex | 0.025 (0.216) | 9.1E-01 | 9.7E-01 |
| | race | -0.172 (0.279) | 5.4E-01 | 7.9E-01 |
| | dual use | 0.325 (0.345) | 3.5E-01 | 6.7E-01 |
| | smoking | 0.686 (0.362) | 5.9E-02 | 2.6E-01 |
| | vaping | 0.393 (0.357) | 2.7E-01 | 6.2E-01 |
| | GOLD 1 vs 0 | 1.022 (0.406) | 1.2E-02 | 1.1E-01 |
| | GOLD 2-4 vs 0 | 0.205 (0.267) | 4.4E-01 | 7.5E-01 |
| | GOLD PrISM vs 0 | 0.053 (0.451) | 9.1E-01 | 9.7E-01 |
| IGHD CDR3 Length | age | -0.001 (0.014) | 9.2E-01 | 9.7E-01 |
| | sex | -0.326 (0.21) | 1.2E-01 | 4.0E-01 |
| | race | 0.96 (0.271) | 4.9E-04 | 1.0E-02 |
| | dual use | -0.116 (0.335) | 7.3E-01 | 9.1E-01 |
| | smoking | -0.224 (0.351) | 5.2E-01 | 7.9E-01 |
| | vaping | -0.364 (0.346) | 2.9E-01 | 6.3E-01 |
| | GOLD 1 vs 0 | -0.047 (0.395) | 9.1E-01 | 9.7E-01 |
| | GOLD 2-4 vs 0 | -0.276 (0.261) | 2.9E-01 | 6.3E-01 |

| | | | | |
|---|---|---|---|---|
| | GOLD PrISM vs 0 | 0.249 (0.427) | 5.6E-01 | 7.9E-01 |
| IGHE CDR3 Length | age | -0.235 (0.098) | 1.8E-02 | 1.2E-01 |
| | sex | 1.189 (1.431) | 4.1E-01 | 7.2E-01 |
| | race | -2.378 (1.843) | 2.0E-01 | 5.3E-01 |
| | dual use | -0.384 (2.294) | 8.7E-01 | 9.6E-01 |
| | smoking | 0.41 (2.392) | 8.6E-01 | 9.6E-01 |
| | vaping | 6.755 (2.427) | 6.0E-03 | 7.6E-02 |
| | GOLD 1 vs 0 | 0.744 (2.795) | 7.9E-01 | 9.4E-01 |
| | GOLD 2-4 vs 0 | 2.652 (1.783) | 1.4E-01 | 4.4E-01 |
| | GOLD PrISM vs 0 | -2.652 (3.057) | 3.9E-01 | 7.1E-01 |
| IGHG1 CDR3 Length | age | -0.013 (0.015) | 3.9E-01 | 7.1E-01 |
| | sex | 0.056 (0.222) | 8.0E-01 | 9.4E-01 |
| | race | 1.038 (0.285) | 3.4E-04 | 9.6E-03 |
| | dual use | -0.424 (0.353) | 2.3E-01 | 5.6E-01 |
| | smoking | -0.107 (0.37) | 7.7E-01 | 9.3E-01 |
| | vaping | -0.167 (0.365) | 6.5E-01 | 8.7E-01 |
| | GOLD 1 vs 0 | -0.307 (0.415) | 4.6E-01 | 7.6E-01 |
| | GOLD 2-4 vs 0 | -0.361 (0.273) | 1.9E-01 | 5.2E-01 |
| | GOLD PrISM vs 0 | -0.389 (0.462) | 4.0E-01 | 7.2E-01 |
| IGHG2 CDR3 Length | age | -0.026 (0.016) | 1.0E-01 | 3.7E-01 |
| | sex | -0.121 (0.23) | 6.0E-01 | 8.2E-01 |
| | race | -0.235 (0.296) | 4.3E-01 | 7.4E-01 |
| | dual use | -0.217 (0.366) | 5.5E-01 | 7.9E-01 |
| | smoking | 0.087 (0.384) | 8.2E-01 | 9.6E-01 |
| | vaping | -0.227 (0.378) | 5.5E-01 | 7.9E-01 |
| | GOLD 1 vs 0 | 0.254 (0.431) | 5.6E-01 | 7.9E-01 |
| | GOLD 2-4 vs 0 | -0.358 (0.283) | 2.1E-01 | 5.3E-01 |
| | GOLD PrISM vs 0 | -0.362 (0.479) | 4.5E-01 | 7.6E-01 |

| | | | | |
|---|---|---|---|---|
| IGHG3 CDR3 Length | age | 0.017 (0.025) | 4.9E-01 | 7.9E-01 |
| | sex | 0.165 (0.364) | 6.5E-01 | 8.7E-01 |
| | race | 0.015 (0.47) | 9.7E-01 | 9.9E-01 |
| | dual use | 0.936 (0.579) | 1.1E-01 | 3.8E-01 |
| | smoking | 0.124 (0.606) | 8.4E-01 | 9.6E-01 |
| | vaping | 0.679 (0.597) | 2.6E-01 | 6.0E-01 |
| | GOLD 1 vs 0 | -0.688 (0.684) | 3.2E-01 | 6.5E-01 |
| | GOLD 2-4 vs 0 | -0.696 (0.45) | 1.2E-01 | 4.0E-01 |
| | GOLD PrISM vs 0 | 0.193 (0.739) | 7.9E-01 | 9.4E-01 |
| IGHM CDR3 Length | age | -0.002 (0.013) | 8.9E-01 | 9.7E-01 |
| | sex | -0.629 (0.185) | 8.1E-04 | 1.6E-02 |
| | race | 0.699 (0.238) | 3.7E-03 | 5.5E-02 |
| | dual use | 0.091 (0.297) | 7.6E-01 | 9.2E-01 |
| | smoking | -0.041 (0.31) | 8.9E-01 | 9.7E-01 |
| | vaping | -0.367 (0.307) | 2.3E-01 | 5.6E-01 |
| | GOLD 1 vs 0 | 0.175 (0.355) | 6.2E-01 | 8.4E-01 |
| | GOLD 2-4 vs 0 | -0.269 (0.229) | 2.4E-01 | 5.8E-01 |
| | GOLD PrISM vs 0 | 0.484 (0.375) | 2.0E-01 | 5.3E-01 |

CDR3 length is the length of the CDR3 sequence in nucleotides. Sex coded as (0=female, 1=male). Race coded as (0=self reported Black race, 1=self reported White race). PrISM - Preserved ratio impaired spirometry.

*Table E6*. **Significant BCR associations to vaping, smoking, and dual use adjusting for socioeconomic status.**

| BCR measure | Dual Use | | | Cigarette smoking | | | Vaping | | |
|---|---|---|---|---|---|---|---|---|---|
| | beta (se) | P-value | q-value | beta (se) | P-value | q-value | beta (se) | P-value | q-value |
| IgA1 usage | 0.101 (0.024) | 3.74E-05 | 2.4E-03 | 0.06 (0.025) | 0.020 | 0.14 | 0.051 (0.025) | 0.038 | 0.19 |
| IgA2 usage | 0.061 (0.011) | 2.26E-07 | 5.7E-05 | 0.033 (0.012) | 0.007 | 0.10 | 0.027 (0.012) | 0.024 | 0.15 |
| IgM usage | -0.185 (0.039) | 4.19E-06 | 5.3E-04 | -0.111 (0.041) | 0.008 | 0.10 | -0.088 (0.04) | 0.029 | 0.16 |
| class switching proportion | 0.119 (0.027) | 2.22E-05 | 1.9E-03 | 0.064 (0.029) | 0.029 | 0.16 | 0.052 (0.028) | 0.066 | 0.32 |
| IGHV1.18.01 usage | 0.005 (0.001) | 4.6E-04 | 0.01 | 0.004 (0.002) | 0.015 | 0.12 | 0.004 (0.002) | 0.014 | 0.12 |
| IGHV3.7.01 usage | 0.006 (0.002) | 5.0E-04 | 0.01 | 0.003 (0.002) | 0.110 | 0.40 | 0.002 (0.002) | 0.365 | 0.71 |
| IGHV5.51.01 usage | 0.009 (0.002) | 8.26E-05 | 4.2E-03 | 0.005 (0.002) | 0.025 | 0.16 | 0.006 (0.002) | 0.012 | 0.12 |
| IgE CDR3 length | -0.193 (2.27) | 0.93 | 0.98 | -1.148 (2.416) | 0.636 | 0.88 | 7.262 (2.372) | 0.003 | 0.05 |

Results from regression model with the following covariates - age, race, sex, smoking/vaping status, GOLD stage, cigarette pack-years, income level, and regional socioecomic deprivation index. Count values are log2 of unique BCR RNA sequence count. Usage is the proportion of all BCR RNA sequences falling into either a specific isotype or v-segment category (calculated separately for isotypes and v-segments). Class switching proportion is the proportion of all BCR RNA sequences that belong to IgA, IgG, or IgE isotypes and have evidence of somatic hypermutation (>1 mutation relative to the IMGT reference database). CDR3 length is the length of the CDR3 sequence in nucleotides.

*Table E7*. **BCR-seq associations comparing never to former smokers.**

| BCR-seq measure | beta | se | p-value |
|---|---|---|---|
| IGHA1 expression | 0.02446 | 0.33579 | 0.94 |
| IGHA2 expression | -0.00339 | 0.34872 | 0.99 |
| IGHD expression | -0.02141 | 0.26909 | 0.94 |
| IGHE expression | -0.05830 | 0.43340 | 0.89 |
| IGHG1 expression | 0.23209 | 0.28251 | 0.41 |
| IGHG2 expression | -0.20065 | 0.30265 | 0.51 |
| IGHG3 expression | -0.14991 | 0.31076 | 0.63 |
| IGHM expression | -0.12241 | 0.25737 | 0.63 |
| IGHA1 usage | 0.01734 | 0.02803 | 0.54 |
| IGHA2 usage | 0.00311 | 0.01334 | 0.82 |
| IGHD usage | 0.00229 | 0.01398 | 0.87 |
| IGHE usage | 0.00005 | 0.00015 | 0.73 |
| IGHG1 usage | 0.01125 | 0.00887 | 0.21 |
| IGHG2 usage | -0.00570 | 0.00657 | 0.39 |
| IGHG3 usage | 0.00152 | 0.00289 | 0.60 |
| IGHM usage | -0.01561 | 0.04578 | 0.73 |
| class switching | 0.00786 | 0.03205 | 0.81 |
| IGHV1.18.01 usage | 0.00007 | 0.00170 | 0.97 |
| IGHV3.7.01 usage | -0.00178 | 0.00192 | 0.35 |
| IGHV5.51.01 usage | 0.00134 | 0.00260 | 0.61 |
| IGHA1 cdr3 length | -0.14497 | 0.35177 | 0.68 |
| IGHA2 cdr3 length | 0.58270 | 0.41113 | 0.16 |
| IGHD cdr3 length | 0.28801 | 0.40174 | 0.47 |
| IGHE cdr3 length | 9.50712 | 2.78003 | 0.001 |
| IGHG1 cdr3 length | -0.02448 | 0.42131 | 0.95 |
| IGHG2 cdr3 length | 0.50042 | 0.43670 | 0.25 |

| | | | |
|---|---|---|---|
| IGHG3 cdr3 length | 1.56373 | 0.69327 | 0.03 |
| IGHM cdr3 length | 0.27231 | 0.35416 | 0.44 |

Count values are log2 of unique BCR RNA sequence count. Usage is the proportion of all BCR RNA sequences falling into either a specific isotype or v-segment category (calculated separately for isotypes and v-segments). Class switching proportion is the proportion of all BCR RNA sequences that belong to IgA, IgG, or IgE isotypes and have evidence of somatic hypermutation (>1 mutation relative to the IMGT reference database). CDR3 length is the length of the CDR3 sequence in nucleotides.

*Table E8*. **BCR-seq associations to self-reported race adjusting for income level and national area deprivation index**

| BCR-seq measure | beta (se) | p-value | q-value |
|---|---|---|---|
| IGHA1 expression | -0.241 (0.246) | 0.3291 | 0.70 |
| IGHA2 expression | -0.162 (0.259) | 0.5316 | 0.81 |
| IGHD expression | -0.056 (0.196) | 0.7740 | 0.92 |
| IGHE expression | 0.07 (0.318) | 0.8266 | 0.94 |
| IGHG1 expression | -0.576 (0.208) | 0.0062 | 0.10 |
| IGHG2 expression | -0.412 (0.224) | 0.0679 | 0.32 |
| IGHG3 expression | -0.719 (0.231) | 0.0022 | 0.04 |
| IGHM expression | 0.06 (0.186) | 0.7469 | 0.90 |
| IGHA1 usage | -0.022 (0.02) | 0.2926 | 0.65 |
| IGHA2 usage | 0 (0.01) | 0.9665 | 0.98 |
| IGHD usage | -0.001 (0.01) | 0.9152 | 0.97 |
| IGHE usage | 0 (0) | 0.1327 | 0.44 |
| IGHG1 usage | -0.023 (0.006) | 0.0004 | 0.01 |
| IGHG2 usage | -0.009 (0.005) | 0.0768 | 0.34 |
| IGHG3 usage | -0.008 (0.002) | 0.0003 | 0.01 |
| IGHM usage | 0.071 (0.033) | 0.0349 | 0.19 |
| class switching | -0.053 (0.023) | 0.0253 | 0.16 |
| IGHV1.18.01 usage | -0.002 (0.001) | 0.0845 | 0.34 |
| IGHV3.7.01 usage | -0.004 (0.001) | 0.0096 | 0.11 |
| IGHV5.51.01 usage | 0 (0.002) | 0.8191 | 0.94 |
| IGHA1 cdr3 length | 0.313 (0.258) | 0.2255 | 0.59 |
| IGHA2 cdr3 length | -0.222 (0.305) | 0.4679 | 0.76 |
| IGHD cdr3 length | 1.033 (0.293) | 0.0005 | 0.01 |
| IGHE cdr3 length | -1.729 (1.951) | 0.3769 | 0.71 |
| IGHG1 cdr3 length | 1.064 (0.311) | 0.0008 | 0.02 |
| IGHG2 cdr3 length | -0.206 (0.319) | 0.5199 | 0.80 |
| IGHG3 cdr3 length | 0.186 (0.512) | 0.7163 | 0.90 |

| | | | |
|---|---|---|---|
| IGHM cdr3 length | 0.649 (0.259) | 0.0131 | 0.12 |

Count values are log2 of unique BCR RNA sequence count. Usage is the proportion of all BCR RNA sequences falling into either a specific isotype or v-segment category (calculated separately for isotypes and v-segments). Class switching proportion is the proportion of all BCR RNA sequences that belong to IgA, IgG, or IgE isotypes and have evidence of somatic hypermutation (>1 mutation relative to the IMGT reference database). CDR3 length is the length of the CDR3 sequence in nucleotides.

*Table E9*. **Significant "Univariate" Associations to COPD**

| Dependent variable | Independent variable | beta_se | P-value | q-value |
|---|---|---|---|---|
| IGHV5.51.01 usage | GOLD 234 vs 0 | 0.005 (0.001) | 0.0003 | 0.03 |
| IGHA2 usage | GOLD 234 vs 0 | 0.025 (0.008) | 0.0017 | 0.07 |
| IGHA1 usage | GOLD 234 vs 0 | 0.049 (0.016) | 0.0026 | 0.07 |
| IGHM usage | GOLD 234 vs 0 | -0.081 (0.027) | 0.0032 | 0.07 |
| class switching | GOLD 234 vs 0 | 0.054 (0.019) | 0.0049 | 0.08 |
| IGHM count | GOLD 234 vs 0 | -0.398 (0.144) | 0.0062 | 0.09 |

Associations are from models with the BCR measure as the response adjusting for only GOLD spirometric stage and inhaled corticosteroid use. Count values are log2 of unique BCR RNA sequence count. GOLD stage coded as GOLD 2-4, GOLD 1, and PrISM with GOLD 0 as the reference. Usage is the proportion of all BCR RNA sequences falling into either a specific isotype or v-segment category (calculated separately for isotypes and v-segments). Class switching proportion is the proportion of all BCR RNA sequences that belong to IgA, IgG, or IgE isotypes and have evidence of somatic hypermutation (>1 mutation relative to the IMGT reference database).

*Table E10*. **Significant "Univariate" Associations to Airway Wall Thickness**

| Dependent variable | Independent variable | beta_se | P-value | q-value |
|---|---|---|---|---|
| IGHV5.51.01 usage | Wall Area Thickness | 0.0003 (0.00009) | 0.002 | 0.055 |
| IGHM count | Wall Area Thickness | -0.023 (0.008) | 0.004 | 0.055 |

Associations are from models with the BCR measure as the response adjusting for only Wall Area Thickness, CT scanner model, and inhaled corticosteroid use. Wall area thickness % is measured as the average thickness of segmental airway walls / total airway area. Count values are log2 of unique BCR RNA sequence count. Usage is the proportion of all BCR RNA sequences falling into either a specific isotype or v-segment category (calculated separately for isotypes and v-segments).
